# OmicsPred as a centralised resource for genetic prediction of multi-omic traits

**DOI:** 10.64898/2026.05.15.26353298

**Authors:** Carles Foguet, Laurent Gil, Yu Xu, Sofía Salazar-Magaña, Scott C. Ritchie, Elodie Persyn, Hae Kyung Im, Michael Inouye, Samuel A. Lambert

## Abstract

Genetic prediction of multi-omic data has emerged as a cost-effective alternative to direct omics profiling, particularly useful for identifying molecular features associated with disease susceptibility. However, despite its popularity, multi-omic imputation models are fragmented across studies, hindering findability, accessibility, interoperability and re-use. To address this, we developed OmicsPred (https://www.omicspred.org), a centralised platform for the deposition and dissemination of genetic prediction models of multi-omic traits. OmicsPred unifies the most commonly used molecular imputation models (e.g. from PredictDB) and other published studies totalling 3,339,469 prediction models spanning transcriptomic, proteomic, and metabolomic traits (as of May 2026). Each model is accompanied by metadata describing score development and predictive performance, and distributed in formats compatible with popular analytic tools, such as PGS Catalog Calculator and MetaXcan. To demonstrate the utility of the resource for systematic target discovery, we perform a multi-omic phenome-wide association analysis in Million Veterans Program data.

## Introduction

RNA transcripts, proteins and metabolites are key mediators of complex phenotypes in health and disease, and quantifying their variation is essential to unravelling disease aetiology and the downstream development of therapeutics. In the last decade, there have been major efforts to collect multi-omic data from various organs and tissues^1,2^; however, comprehensive multi-tissue, transcriptomic, proteomic, and metabolomic profiling remains unavailable for most large cohorts due to practical limitations and cost. In contrast, genotype data are now widely available across global biobanks and population studies. Studies of genotyped cohorts with multi-omics data have identified robust QTLs and established that many molecular trait levels are strongly heritable^2,3^. Given the heritability of these traits, genetic prediction of omic traits has emerged as a powerful and cost-effective alternative to direct profiling^4–6^.

Genetic prediction models are trained in datasets with paired genotype and omic data, where first the genetic variants associated with a given molecular feature are identified, then combined into a weighted genetic score that captures the contribution of each genetic variant to each molecular trait. The most popular set of such models is from PredictDB, comprising 49 tissue-specific transcriptome models trained using the GTEx bulk gene expression data^5^. More recently, larger cohorts with multi-omic data have been leveraged to train genetic prediction models for blood transcriptomic, proteomic and metabolomic traits^6–8^. Similar genetic prediction models have also been developed for farm animals^9^, highlighting the broad interest and utility of genetic prediction models across fields. The resulting genetic models can be used to impute individual-level omic traits in large cohorts or biobanks, enabling downstream analysis, such as transcriptome-, proteome- or metabolome-wide association studies (TWAS, PWAS and MWAS, respectively)^6^. Alternatively, these models can be applied directly to genome-wide association study (GWAS) summary statistics to identify molecular traits associated with disease risk and other complex phenotypes (e.g. S-PrediXcan, FUSION)^4,10^, allowing researchers to leverage the vast trove of publicly available GWAS results^11^ without the need for individual-level data.

Despite the widespread use of genetic prediction models for omic traits, it has been a longstanding challenge to make them findable, accessible, interoperable and re-usable in accordance with FAIR Data Principles. For example, there is a lack of standardisation in model files and their metadata, and existing models are often dispersed across supplementary materials or study-specific websites. Resources like the GWAS and Polygenic Score (PGS) catalogues^11,12^ have succeeded in making GWAS summary statistics and phenotype prediction more accessible, but these do not extend to genetic prediction models for molecular traits or integrate with established TWAS, PWAS or MWAS frameworks. To address this gap, we present OmicsPred (https://www.omicspred.org/), a centralised resource for hosting and disseminating genetic prediction models of multi-omic traits and their metadata. We provide an overview of the key features of OmicsPred and demonstrate its applications by performing a phenome-wide association analysis in Million Veterans Program data^13^.

### The OmicsPred resource

OmicsPred functions as a centralised repository for hosting and disseminating genetic prediction models of multi-omic traits, modelled on FAIR principles of scientific data management. Key metadata is provided for each score (**Figure 1**), including training set characteristics (e.g., cohort name, sample size and genetic ancestry) and predictive performance (R^2^) in held-out training data or external cohorts. The molecular entity each score predicts, gene, protein, or metabolite, are mapped to appropriate reference databases (Ensembl, Uniprot, or ChEBI respectively), along with EBI’s experimental factor ontology (EFO) for tissue terms. To facilitate biological interpretation, molecular features are mapped to Reactome biological pathways, allowing users to query genes, proteins, and metabolite models at the pathway level (**Supplemental Note 1**). Scoring files are harmonised to facilitate interoperability, and can be downloaded in both PGSC-Calculator^12^ and MetaXcan^10^ compatible formats. Scores derived from a single publication that share a common platform, tissue, and training methodology are aggregated into score datasets to provide a coherent unit for browsing and downstream analyses. Hosted scores can be browsed and searched by dataset, molecular entity, pathway, omics-layer/platform, tissue or publication via a web interface, and downloaded or programmatically queried via REST API (**Figure 1, Supplemental Note 2**).

**Figure 1.**
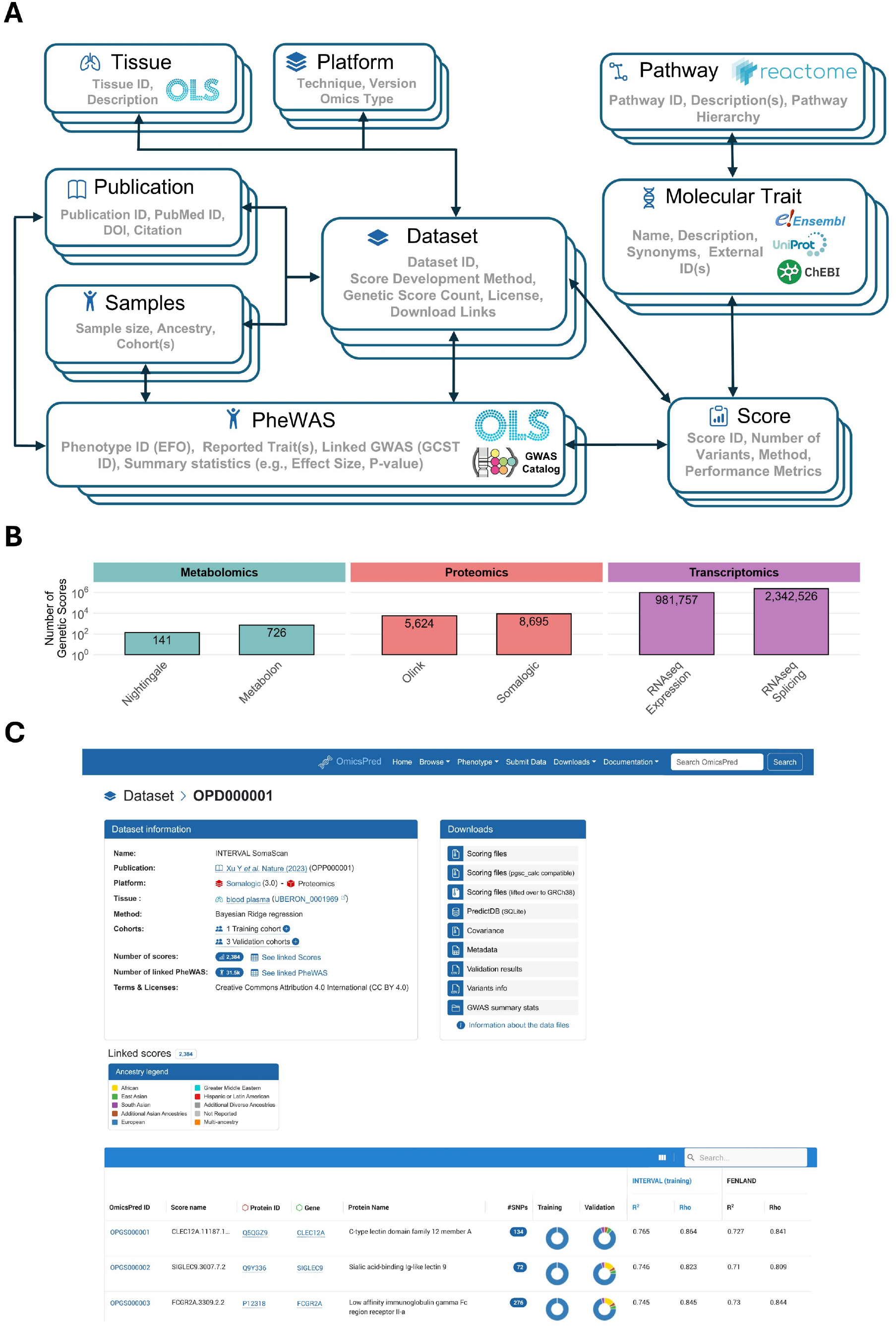
Overview of the OmicsPred resource and interface. **(A)** Schematic representation of the metadata structure in OmicsPred. **(B)** Summary of the number of genetic scores (presented on a log scale) included in OmicsPred as of May 2026, stratified by omic-type and platform. **(C)** Example of a Dataset page for OPD000001 (https://www.omicspred.org/dataset/OPD000001). Key Dataset metadata, like source publication, platform, training method, and training/validation cohorts, are indicated. Scores that are part of the dataset are listed below (here, 3 out of 2384 are shown) along with key metadata such as gene and protein identifiers, training and validation sample descriptors (including ancestry), and score predictive performance. Download links are provided for the genetics scores in both PGSC Calculator and MetaXcan compatible formats, along with links to the complete metadata for the dataset. A link to the PheWAS results page for the dataset is also provided.

As of May 2026, OmicsPred hosts 3,339,469 prediction models spanning transcriptomic, proteomic, and metabolomic features (**Figure 1**). The majority of scores are from the GTExV8 Elastic Net and MASHR PredictDB datasets^5^, enabling the imputation of RNA-seq gene expression and splicing events across 49 human tissues. For diversity across omic platforms, the repository includes 17,227 models trained in the INTERVAL cohort for the prediction of blood transcriptomic, proteomic (profiled using the antibody-based Olink and the aptamer-based SomaLogic platforms), and metabolomic traits (profiled using the NMR-based Nightingale and MS-based Metabolon platforms)^6^. Furthermore, OmicsPred also includes SomaLogic-based plasma proteomics prediction models trained in diverse genetic ancestries, namely 3,578 models trained in the Trans-omics for Precision Medicine (TOPMed) Multi-omics pilot study^7^ and 2,733 models trained in the Atherosclerosis Risk in Communities (ARIC) study^8^. Finally, OmicsPred includes Olink proteomics scores trained on the latest UKB Olink data release (2,612 genetic prediction models trained in a subset of 34,557 UKB participants of European genetic ancestries and 2,704 models trained in a multi-ancestry subset of 52,315 UKB participants).

OmicsPred supports phenome-wide association studies (PheWAS) as a source of hypothesis-generation for experimental follow-up by providing a comprehensive catalogue of associations between genetically predicted molecular traits and disease outcomes. PheWAS results can be queried by prediction score, dataset, publication or phenotype. To facilitate cross-study comparability, phenotypes are mapped to reference ontology terms using EFO (e.g. Mondo Disease Ontology). Study metadata is also provided, including testing cohort, genetic ancestry, sample sizes and the statistical model used. For analyses performed using GWAS summary statistics (e.g. with S-PrediXcan^10^), links to the corresponding studies in the GWAS Catalog^11^ are included.

Researchers are encouraged to submit new prediction models via an online form (www.omicspred.org/submit) to facilitate independent replication as well as maximise the impact, findability and reusability of their models. New submissions require score model files, information on the dataset used to train the scores (e.g. platform, source tissue, sample size, cohort characteristics, computation method), performance metrics and a corresponding publication that should be cited when using the scores. PheWAS summary stats for the submitted scores can also be provided. The OmicsPred team will assist and guide the researchers through the submission process, ensuring the submitted scores and their metadata are properly formatted and meet the inclusion criteria of the resource (**Supplemental Note 3**).

### Phenome-wide insights using OmicsPred

Molecular imputation can be a powerful tool for target prioritisation^4–6^. Here, we show how data in OmicsPred can be used to systematically identify gene expression (TWAS) and protein (PWAS) associations across the human phenome using the Million Veterans Program (MVP) cohort, one of the world’s largest multi-ancestry studies^13^. To conduct our PheWAS, we utilised harmonised GWAS summary statistics for up to 1,233 health outcomes (PheCodes) in African (AFR), Admixed American (AMR), and European (EUR) genetic ancestry groups^11,13^. To prioritise genes and proteins we used OmicsPred to identify and download relevant imputation models, focusing on blood and plasma, for each analysed ancestry^5–8^ and performed TWAS and PWAS using MetaXcan.^10^ In total, we identified >190,000 significant associations (FDR<0.05) between imputed transcript and protein traits and PheCode outcomes (∼46M tests; **Table S1**).

Leveraging the cross-dataset score mapping provided by OmicsPred, we identified numerous instances in which gene products were significantly associated with disease risk across omic layers, platforms and ancestries. These include well-established associations such as *PCSK9* with coronary artery disease, *APOE* with hyperlipidemia and *CFH* with age-related macular degeneration (**Figure 2**). Notably, one of the most consistent cross-omic and cross-ancestry signals was the association between *AGER* and hypothyroidism risk. This association was significant in 7 of the 9 analysed datasets and is consistent with the experimental evidence that AGER levels in blood are dysregulated in hypothyroidism^14^. Using the Reactome biological pathway mappings provided by OmicsPred, we were able to ascertain that AGER is a receptor that promotes NF-κB complex activation upon ligand binding (Reactome Pathway R-HSA-445989), consistent with the key role of NF-κB signalling in thyroid physiology and autoimmunity^15^. The linkage between OmicsPred and Reactome pathways enabled identification of additional OmicsPred scores mapping to the same pathway that were also significantly associated with hypothyroidism, including scores mapping to *NFKB2, NFKBIA* and *APP* (itself a ligand of AGER).

**Figure 2.**
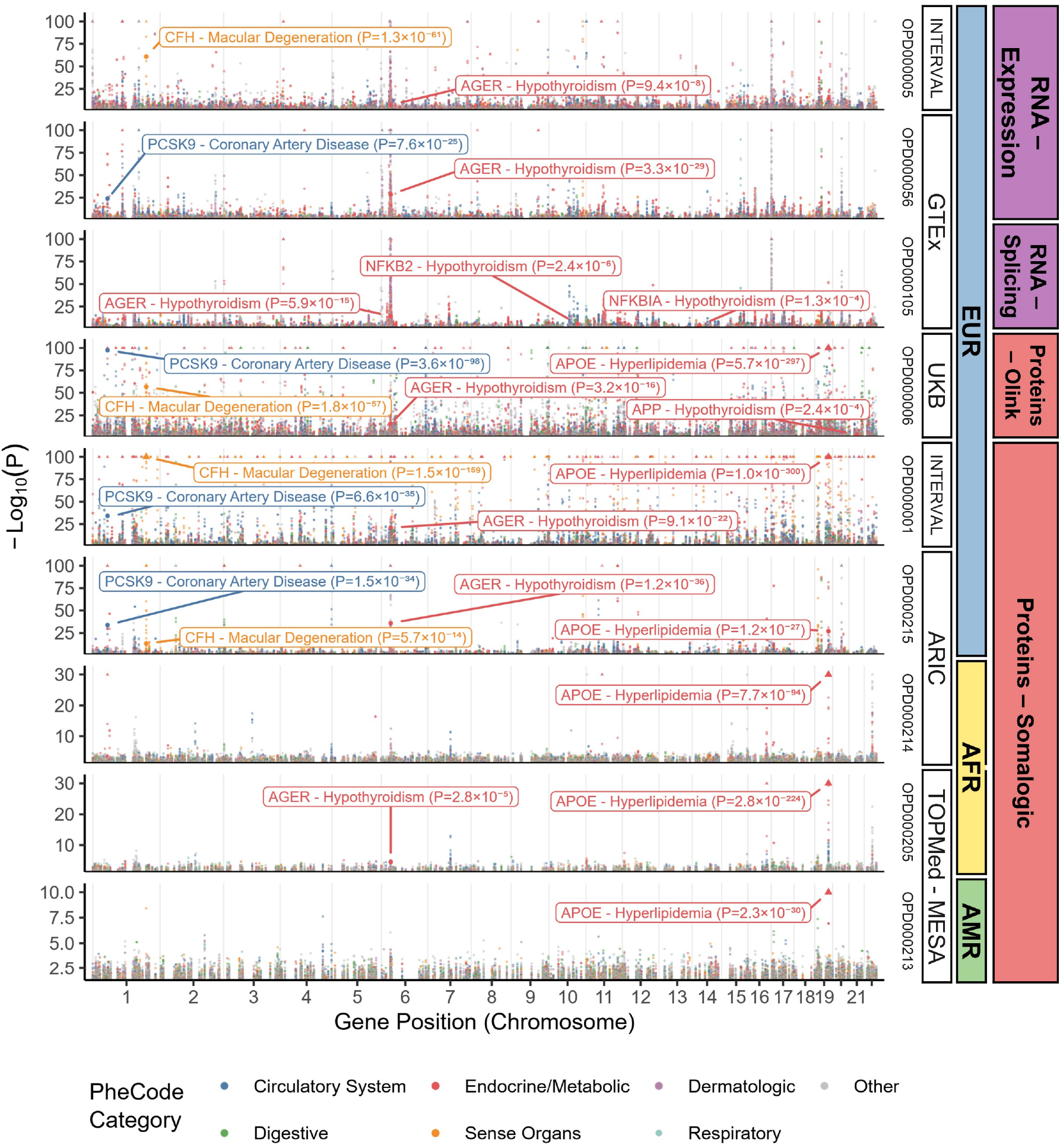
Multi-omic PheWAS in the MVP cohort. Associations between genetically predicted omic features and PheWAS phenotypes (PheCodes) were estimated using S-PrediXcan (MetaXcan) with GWAS summary statistics from the MVP cohort as input. Each dataset was annotated according to the data modality and platform evaluated in the MVP ancestry subset most closely matching the dataset’s training cohort (right side of plot). Associations are plotted according to statistical significance (-log_10_ p-value), coloured by PheCode category; upward-pointing triangles indicate associations exceeding the y-axis range. Associations discussed in the manuscript are highlighted.

The MVP PheWAS results are incorporated into OmicsPred together with the results from a previously published UK Biobank PheWAS using PGSC-Calculator imputed molecular scores^6^. OmicsPred maps identified associations to scores, proteins and genes within the context of biological pathways across globally diverse biobanks, thus facilitating target validation and hypothesis generation for downstream studies.

## Conclusion & future developments

OmicsPred provides a unified FAIR repository for multi-omic genetic prediction models, together with a catalogue of associations between genetically-predicted molecular traits and health outcomes, facilitating broad applications in biomedical research. Linkage to external resources such as Reactome and the GWAS Catalog, together with ontology-based mappings for tissues and phenotypes further improves the annotations and interpretation of association results. While OmicsPred is biased towards scores developed in predominantly European ancestry cohorts, the development of omic imputation models in global populations is a growing area of research, and we are committed to prioritising inclusion of such studies^7,8^ to maximise the diversity and equitable application of genetic scores as data becomes available.

We encourage researchers to submit newly developed genetic prediction models and associated PheWAS results to OmicsPred, to increase the findability and reusability of their work by the broader scientific community (www.omicspred.org/submit). We anticipate that OmicsPred will continue to grow in scale and diversity through new submissions, expanding to include models trained across additional tissues, ancestries and molecular platforms (e.g. mass spectrometry-based proteomics). With growing research using genetic prediction of multi-omics in non-human species (e.g. farm animals^9^), we plan to realise the synergies of linking these data in a pan-species database. Future plans also include new features to support advanced analyses, such as cis-partitioning of scores (for causal association testing), better linkage of metabolite data to gene regions (e.g. metabolising enzymes), and inclusion of protein complex data. Taken together, OmicsPed now provides an integrated database to support the use of imputed multi-omics in biomedical research.

## Data Availability

Data is available through www.omicspred.org

## Acknowledgements

We thank the authors of publications in OmicsPred for making their data open-access so that it can be made available to the community and indexable in our database, and those who responded to our inquiries and requests for data.

This work was supported by core funding from the British Heart Foundation (RG/F/23/110103), NIHR Cambridge Biomedical Research Centre (NIHR203312) [*], BHF Chair Award (CH/12/2/29428), Cambridge BHF Centre of Research Excellence (RE/24/130011), and by Health Data Research UK (HDRUK2023.0028), which is funded by the UK Medical Research Council, Engineering and Physical Sciences Research Council, Economic and Social Research Council, Department of Health and Social Care (England), Chief Scientist Office of the Scottish Government Health and Social Care Directorates, Health and Social Care Research and Development Division (Welsh Government), Public Health Agency (Northern Ireland), British Heart Foundation and the Wellcome Trust. S.C.R. is funded by the BHF Cambridge Centre for Research Excellence (RE/24/130011); M.I. is supported by the Munz Chair of Cardiovascular Prediction and Prevention and the NIHR Cambridge Biomedical Research Centre (NIHR203312) [*] as well as by the UK Economic and Social Research Council (ES/T013192/1).

