## Supplemental Notes 1-3, Table S1, Figures S1-6 for "OmicsPred as a centralised resource for genetic prediction of multi-omic traits"

### Supplemental Material

**Supplemental Notes** **1**

1. OmicsPred scoring file and metadata description 1

Performance Metrics (used to describe training and testing performance for scores) 6

2. OmicsPred data access and implementation 8

3. Inclusion Criteria for OmicsPred 8

**Supplemental Tables** **9**

**Supplemental Figures** **11**

**Supplemental References** **16**

### Supplemental Notes

#### 1. OmicsPred scoring file and metadata description

The goal of the OmicsPred resource is to make the genetic scores used for molecular imputation and association studies findable, accessible, interoperable and re-usable in accordance with FAIR Data Principles[1].

The OmicsPred scoring file format is based on the scoring file format of the PGS Catalog[2] (https://www.pgscatalog.org/downloads/#dl_ftp_scoring). Each scoring file is labelled by its OPGS ID (e.g. OPGS000001.txt). The scoring files are zipped by dataset.

The header of the Genetic Scoring files are slightly different from the one in PGS Catalog (highlighted with '#' below):

```
# OmicsPred ID = OPGS identifier, e.g. 'OPGS000001'
# PGS Name = Genetic Score name, e.g. 'OID21313'
# Trait type = omics type (e.g. proteomics)
# Measurement tissue = tissue name, e.g. venous blood
(UBERON_0013756)
# Measurement platform = platform, e.g. Olink
# Reported Trait = molecular trait, e.g. 'Dickkopf-like protein 1'
# Original Genome Build = Genome build/assembly, e.g. 'GRCh38'
# Number of Variants = Number of variants listed in the Genetic
Score
# Note = extra information, e.g. 'Model extracted as-is from
PredictDB.org'
# Citation = Information about the publication
# License = License and terms of the Genetic Score
use/distribution
rsID  chr_name  chr_position  effect_allele  reference_allele
```

```
effect_weight
…
```

The columns defining the data format itself are listed below:

| Column Header | Field Name | Field Description | Is Mandatory? |
|---|---|---|---|
| *rsID* | dbSNP Accession ID (rsID) | Unique identifier of the variant in dbSNP | **Yes** - Each Genetic Score file must have either an *rsID* column or both a *chr_name* and *chr_position* column to identify the variant. |
| *chr_name* | Location - Chromosome | Chromosome associated with the variant | |
| *chr_position* | Location - Base pair position within the Chromosome | Chromosomal position associated with the variant | |
| *effect_allele* | Effect allele | The allele that's dosage is counted (e.g. {0, 1, 2}) and multiplied by the variant's weight (*effect_weight*) when calculating score. The effect allele is also known as the 'risk allele' | **Yes** |
| *other_allele* | Other allele | The other allele(s) at the loci. | **Suggested** - most software requires this for the calculation of scores and matching of the variants to existing genotype data |
| *effect_weight* | Variant weight | Value of the effect that is multiplied by the dosage of the effect allele (*effect_allele*) when calculating the score | **Yes** |

This table describes the data items that can be captured for each of the data objects in OmicsPred

| OmicsPred Data Objects | Data Items | Description | Comments |
|---|---|---|---|
| **Publication** (Identified by OPP ID) | OmicsPred Publication ID (OPP) | Unique identifier created for the publication entries in OmicsPred. | This information is extracted and annotated according to EuropePMC [3]. |
| | PubMed ID (PMID) | PubMed Identification number. | |
| | Digital Object Identifier (doi) | The doi of the publication. | |
| | Title | Title of the publication. | |
| | Author(s) | List of publication authors, the first author is also extracted for a shorter display. | |
| | Journal | The name of the publication source. | |
| | Publication Date | Date of publication (with respect to the PMID or doi). | |
| **Platform** | Platform name | Name of the platform. | |
| | Platform full name | Full name of the platform. | |
| | Technic | Short description of the technic used on the platform. | |
| | Type | Omics type detected by the platform. | |
| | Version | Platform version (if available). | |
| **Tissue** | Identifier | External identifier from an ontology (e.g. Uberon) | This information is extracted and annotated according to Experimental Factor Ontology (EFO)[4] using the Ontology Lookup Service (OLS)[5]. |
| | Label | Tissue short name/label | |
| | Description | Detailed description of the Tissue | |
| | URL | External URL to the ontology entry | |
| **Pathway** | External Identifier | Pathway external identifier (e.g. R-HSA-156582 from Reactome) | Linked to Reactome [6] |

| | | | |
|---|---|---|---|
| | External Source | Pathway external source (e.g. Reactome) | |
| | Parent External Identifier | List of parent Pathway external ID(s) | |
| | Top Level | Flag to indicate if the Pathway is top-level or not in Reactome | |
| | Super Pathways | List of top-level parent Pathway | |
| **Molecular Trait** (Gene, Protein, Metabolite) | Name | Name of the Molecular Trait. | |
| | External Identifier | Molecular Trait external identifier (e.g. P16581 from UniProt). | Linked to resources such as Ensembl [7], UniProt [8] and ChEBI [9] |
| | External Source | Molecular Trait external source (e.g. Ensembl, UniProt, ChEBI). | |
| | Synonyms | List of Molecular Trait synonyms. | |
| | External References | List of Molecular Trait external references. | |
| | Biotype | Molecular trait biotype. | Only for Gene |
| | Retired gene model | Indicate if the Molecular Trait entry has been retired/removed for the external source. | Only for Gene |
| | Gene | Associated Gene. | Only for Protein |
| | Pathways | Associated Pathways. | Using mapping provided by Reactome [6] |
| **Dataset** (Identified by OPD ID) | OmicsPred Dataset ID (OPD) | Unique identifier created for the dataset entries in OmicsPred. | |
| | Dataset Name | Name of the dataset (if available). | |
| | Publication | Associated Publication. | |
| | Platform | Associated Platform. | |
| | Omics Type | Data type detected by the platform (e.g. gene, protein, metabolite). | |
| | Genetic Score Development Method | The name or description of the method or computational | |

| | | | |
|---|---|---|---|
| | | algorithm used to develop the Genetic Scores of this dataset. | |
| | Genetic Score Count | Number of Genetic Scores associated with the dataset | |
| | Tissue | Biological tissue collected to be analyzed on the platform | |
| | Training Sample(s) | Set of Samples used to create and train the Genetic Scores in the dataset | |
| | Validation Sample(s) | Set of Samples used to validate the Genetic Scores in the dataset | |
| | Data files URLs | JSON structure listing the URLs of the different types of datafiles available for download. | |
| | License/Terms of Use | License/Terms of Use that applies to the Genetic Scores of the Dataset. | |
| **Genetic Score** (Identified by OPGS ID) | OmicsPred ID | Unique identifier created for the genetic score in OmicsPred. | |
| | OmicsPred name | Name used by the author to refer to the genetic score before an OmicsPred identifier has been assigned. | |
| | Reported Molecular Trait | Molecular trait (Gene, Protein, Metabolite, ...) identifier and/or name as reported by the author. | |
| | Original Genome Build | The version of the genome that the variants present in the Genetic Score are associated with. | |
| | Number of Variants | Number of variants used to calculate the Genetic Score. In the future this will include a more detailed description of the types of variants present. | |
| | Genetic Score Development Method | The name or description of the method or computational algorithm used to develop the Genetic Score. | |
| | Molecular Traits | Molecular Traits linked to the Genetic Score: Gene, Transcript, Protein, Metabolite. | Linked to resources such as Ensembl [7], UniProt [8] and ChEBI [9] |

| | | |
|---|---|---|
| | Ancestry distribution | Distribution of the ancestries in the training and validation of the Genetic Score. |
| | Dataset | Associated Dataset |
| | License/Terms of Use | License/Terms of Use that applies to the Genetic Score. |
| **Cohort** | Cohort name | Cohort short name. |
| | Cohort full name | Full name of the cohort. |
| | URL | Link to the cohort/study website. |
| **Sample** | Sample Number | Number of individuals included in the sample. |
| | Sample Cases | Number of individual cases in the sample. |
| | Sample Controls | Number of individual controls in the sample. |
| | Sample Male Percent | Percentage of male individuals in the sample |
| | Broad Ancestry Category | Author reported ancestry is mapped to the best matching ancestry category from the NHGRI-EBI GWAS Catalog framework (Table)[10]. |
| | Cohort(s) | List of Cohorts used to create the Sample. |
| **Performance Metrics** (used to describe training and testing performance for scores) | Genetic Score | Associated Genetic Score |
| | Dataset | Associated Dataset (includes Publication, Platform and Tissue) |
| | Sample | Associated Sample |
| | Cohort label | Shortcut to retrieve the cohort short name/label |
| | Metric value | A list of metrics used to evaluate the performance of the Genetic Score within a Sample/Cohort<br>• **Name**: Name of the metric method (e.g. Proportion of the variance explained). |

| | | | |
|---|---|---|---|
| | | • **Name short**: Shorter name of the metric method (e.g. R2).<br>• **Type**: Type of metric (e.g. Pearson's correlation).<br>• **Estimate value**: Metric estimate value.<br>• **P-value**: P-value associated with the Metric. | |
| **Phenotype** (used to organise PheWAS data) | Identifier | External identifier of the Phenotype. | This information is extracted and annotated according to Experimental Factor Ontology (EFO)[4] using the Ontology Lookup Service (OLS)[5]. |
| | Name | Phenotype name/label. | |
| | Description | Detailed description of the Phenotype | |
| | Category | Phenotype category. | |
| | Source | External source of the Phenotype (e.g. EFO). | |
| | Mapped Phecode | List of mapped PheCode IDs | |
| **PheWAS Results** | Score | Associated Score | |
| | Phenotypes | Associated Phenotypes | |
| | Sample | Associated Sample (control/cases, percentage male participants). | |
| | Dataset | Associated Dataset | |
| | Study type | Type of study (imputed or summary) | |
| | Trait reported | Reported phenotype | |
| | Ancestry distribution | Distribution of the ancestries in the Score PheWAS. | |
| | FDR | False Discovery Rate-adjusted P-value (<0.5). | |
| | Other data | • $R^2$<br>• HR: Hazard Ratio with confidence interval<br>• Z-score: Standard score | |

| | | |
|---|---|---|
| | | • P-value<br>• Bonferroni<br>• Effect size<br>• Var_gene_exp: Gene expression variance |
| | Variants number used | Number of variants from the genetic score used in the PheWAS |
| | Variants fraction found | Fraction of variants used from the genetic score used in the PheWAS |

### 2. OmicsPred data access and implementation

Data in OmicsPred can be currently accessed in the following ways:

- **Downloads** from OmicsPred are provisioned by the Box™ service. All the available downloads are listed on the webpage https://www.omicspred.org/downloads. It includes several types of downloads, grouped by dataset:
  - **Metadata files** (excel spreadsheets) describing all genetic scores in terms of their publication source, detection platform, samples, tissues, phenotypes, molecular traits and related performance metrics.
  - **Genetic Scores** are provided as:
    - **Scoring files** compatible with `pgsc_calc`[2] (https://pgsc-calc.readthedocs.io/en/latest/). The data format is described above and on the PGS Catalog website (https://www.pgscatalog.org/downloads/#dl_ftp_scoring).
    - **Databases in PredictDB format** (https://predictdb.org/) that are compatible with MetaXcan tools[11]. Associated covariance files are also available.
- A **REST API** is also provided to allow programmatic access and querying of OmicsPred metadata, better enabling other applications to be built on top of the resource. Endpoints to retrieve all or individual OmicsPred data objects (Publications, Datasets, Genetic Scores, Platforms, Samples, Molecular Traits, Performance Metrics, PheWAS) are available (details at: https://rest.omicspred.org/).

OmicsPred is also indexed on FAIRsharing.org (doi:10.25504/FAIRsharing.784c89).

Additional bibliographic information for OmicsPred **Publication** objects are retrieved from Europe PMC[3] (e.g. title, authors, journal, publication dates). Additional information for each ontology term (e.g. for the Tissues and Phenotypes objects) from the EFO[4] are obtained using the EMBL-EBI Ontology Lookup Service (OLS)[5].

The OmicsPred website is developed using ReactJS (Vite version 8; https://vite.dev/) and is fed by a private REST API. The two REST APIs (public and private) and the database are developed using the Django framework (version 5.2; https://djangoproject.com) in Python (version 3.13; https://www.python.org) with a PostgreSQL database (version 15; https://www.postgresql.org/). The search functionality is built using ElasticSearch (v7.17; https://www.elastic.co).The website, database, REST API and search index are all deployed on the Google Cloud platform (https://cloud.google.com/). The codebase for OmicsPred can be viewed within our public GitHub repository (https://github.com/OmicsPred), currently provided under an Apache 2.0 License.

### 3. Inclusion Criteria for OmicsPred

For a publication's data to be included in OmicsPred it must contain one of the following:

- **Newly developed genetic scores.** This includes the following information about the score and its predictive ability (evaluated on samples not used to develop the score):
    - Variant information necessary to apply the score to new samples (variant rsID and/or genomic position, weights/effect sizes, effect allele, genome build).
        - *Optional*: covariance matrix for variants to support summary-statistics based analyses (e.g. MetaXcan).
    - Information about score development (computational method, variant selection, relevant parameters).
    - Descriptions of the samples used to develop and evaluate each score .
    - Establishment of analytic validity and a description of each score's predictive performance, typically the proportion of the variance explained ($R^2$).
- **An evaluation of a previously developed OmicsPred score or dataset**, for instance on samples not used for development. The requirements for description would be the same as for the evaluation of a new genetic score.
- **PheWAS results of genetic scores** for organismal phenotypes (e.g. disease traits) can also be accepted, provided submitters can demonstrate impact to downstream users.

A complete description of the metadata captured is described in Supplemental Note 2 and the most up to date version can be found on the website(https://www.omicspred.org/docs). To ensure comprehensiveness of the resource we will actively identify new publications with scores via literature search and contact authors directly to encourage submission to the resource.

### Supplemental Tables

**Supplementary Table 1. Summary of MVP PheWAS analyses and results.** Eight OmicsPred score datasets derived from blood and plasma were evaluated. Association analyses were performed using the S-PrediXcan function implemented in MetaXcan, with MVP GWAS summary statistics for PheCodes traits as input. Each score dataset was analysed only in GWAS with ancestry similar to the training ancestry. The resulting p-values were corrected for multiple testing within each PheCode-dataset combination. Scores with less than 75% of variant match rate to the GWAS summary stats were excluded from analysis.

| Dataset ID | Tissue | Publication | Omic Layer | Platform | Training Cohort (Ancestry) | #Scores | #GWAS Traits | GWAS Ancestry | FDR Significant Associations |
|---|---|---|---|---|---|---|---|---|---|
| OPD000001 | blood plasma | OPP000001 | Proteomics | Somalogic | INTERVAL (European) | 2,384 | 1233 | European | 26,109 |
| OPD000005 | blood | OPP000001 | Transcripo-mics | RNAseq - Expression | INTERVAL (European) | 13,668 | 1233 | European | 39,586 |
| OPD000006 | blood plasma | OPP000002 | Transcripo-mics | Olink | UK Biobank (European) | 2,612 | 1233 | European | 53,669 |
| OPD000056 | venous blood | OPP000003 | Transcripo-mics | RNAseq - Expression | GTEx (Largely European) | 7,252 | 1233 | European | 30,552 |
| OPD000105 | venous blood | OPP000003 | Transcripo-mics | RNAseq - Splicing | GTEx (Largely European) | 8,724 | 1233 | European | 36,442 |
| OPD000205 | blood plasma | OPP000004 | Proteomics | Somalogic | TOPMed - MESA (African) | 557 | 986 | African | 275 |
| OPD000213 | blood plasma | OPP000004 | Proteomics | Somalogic | TOPMed - MESA (Ad Mixed American) | 584 | 719 | Admixed American | 117 |
| OPD000214 | blood plasma | OPP000006 | Proteomics | Somalogic | ARIC (African) | 1359 | 986 | African | 419 |
| OPD000215 | blood plasma | OPP000006 | Proteomics | Somalogic | ARIC (European) | 1310 | 1233 | European | 6969 |

### Supplemental Figures

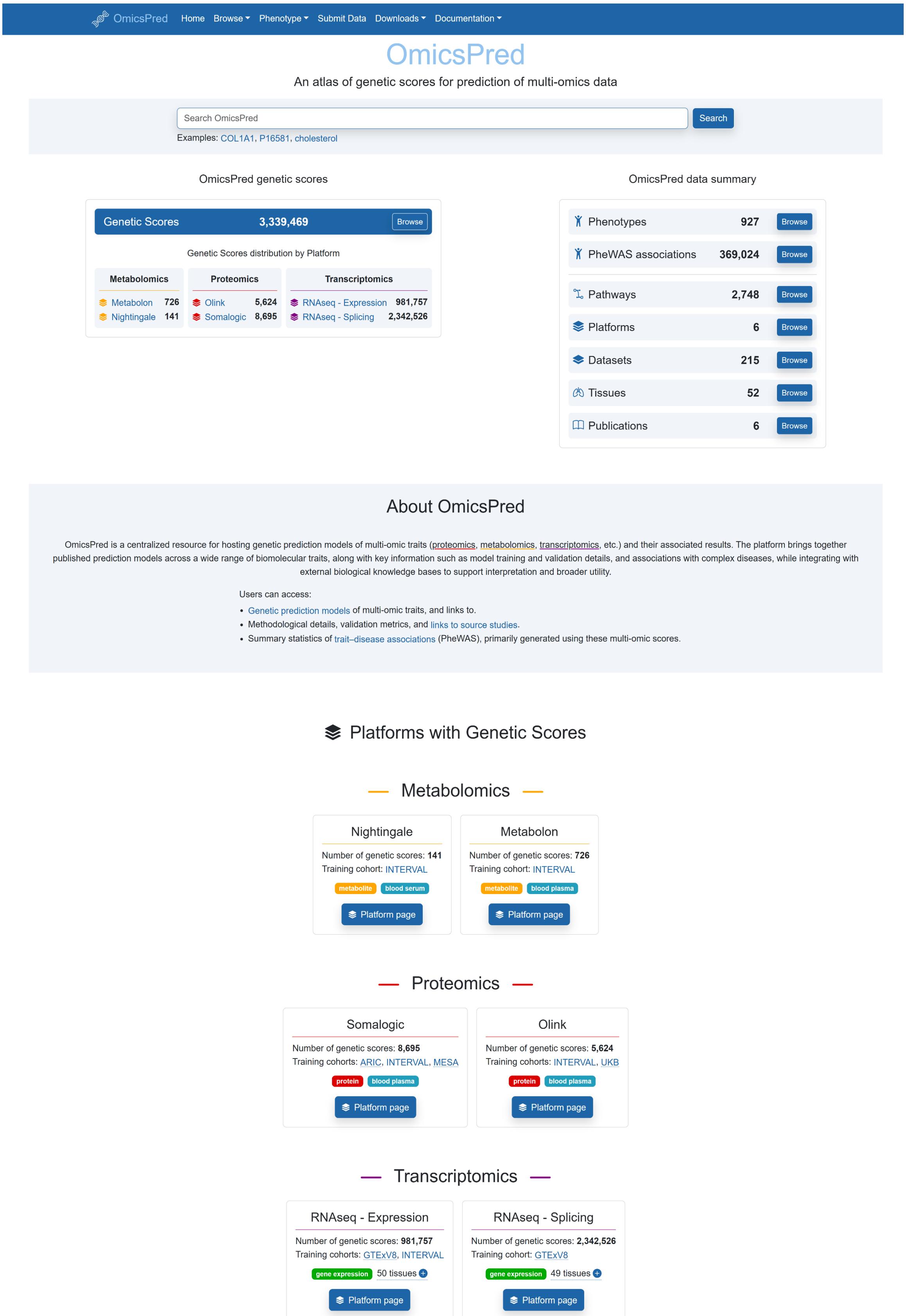

**Figure S1. Screenshot of the OmicsPred landing page (www.omicspred.org).** This page gives users a visual overview of the data indexed in the Catalog.

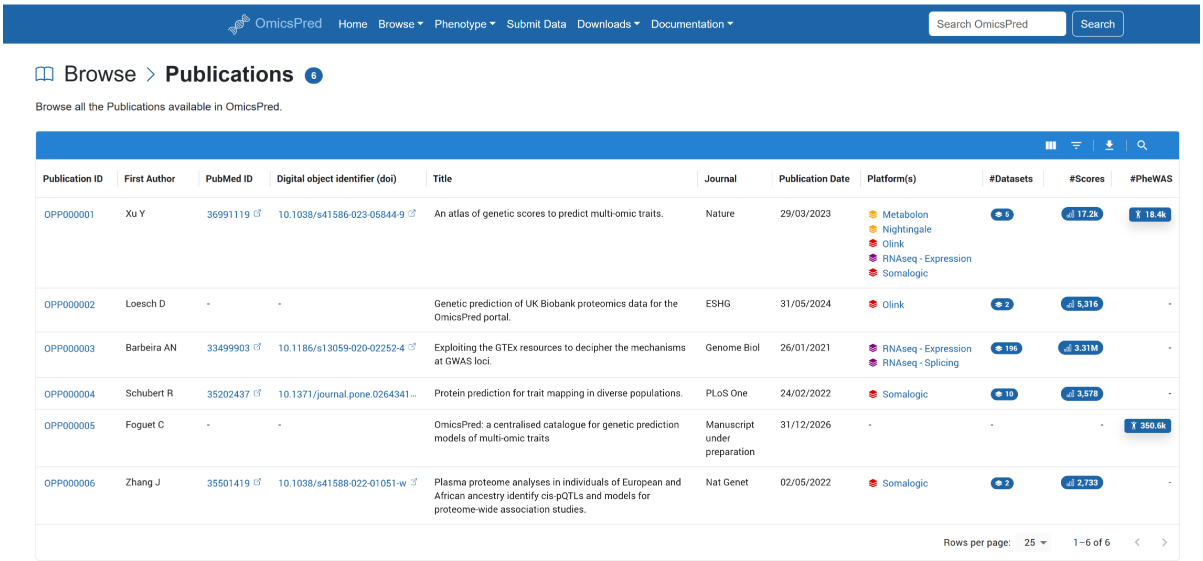

OmicsPred Home Browse Phenotype Submit Data Downloads Documentation Search OmicsPred Search

Browse > Publications 6

Browse all the Publications available in OmicsPred.

| Publication ID | First Author | PubMed ID | Digital object identifier (doi) | Title | Journal | Publication Date | Platform(s) | #Datasets | #Scores | #PheWAS |
|---|---|---|---|---|---|---|---|---|---|---|
| OPP000001 | Xu Y | 36991119 | 10.1038/s41586-023-05844-9 | An atlas of genetic scores to predict multi-omic traits. | Nature | 29/03/2023 | Metabolon<br>Nightingale<br>Olink<br>RNAseq - Expression<br>Somalogic | 5 | 17.2k | 18.4k |
| OPP000002 | Loesch D | - | - | Genetic prediction of UK Biobank proteomics data for the OmicsPred portal. | ESHG | 31/05/2024 | Olink | 2 | 5,316 | - |
| OPP000003 | Barbeira AN | 33499903 | 10.1186/s13059-020-02252-4 | Exploiting the GTEx resources to decipher the mechanisms at GWAS loci. | Genome Biol | 26/01/2021 | RNAseq - Expression<br>RNAseq - Splicing | 196 | 3.31M | - |
| OPP000004 | Schubert R | 35202437 | 10.1371/journal.pone.0264341... | Protein prediction for trait mapping in diverse populations. | PLoS One | 24/02/2022 | Somalogic | 10 | 3,578 | - |
| OPP000005 | Foguet C | - | - | OmicsPred: a centralised catalogue for genetic prediction models of multi-omic traits | Manuscript under preparation | 31/12/2026 | - | - | - | 350.6k |
| OPP000006 | Zhang J | 35501419 | 10.1038/s41588-022-01051-w | Plasma proteome analyses in individuals of European and African ancestry identify cis-pQTLs and models for proteome-wide association studies. | Nat Genet | 02/05/2022 | Somalogic | 2 | 2,733 | - |

Rows per page: 25 1–6 of 6

**Figure S2. Screenshot of the OmicsPred publication page ([https://www.omicspred.org/publications](https://www.omicspred.org/publications)).** This page allows users to browse the list of publications included in the catalog.

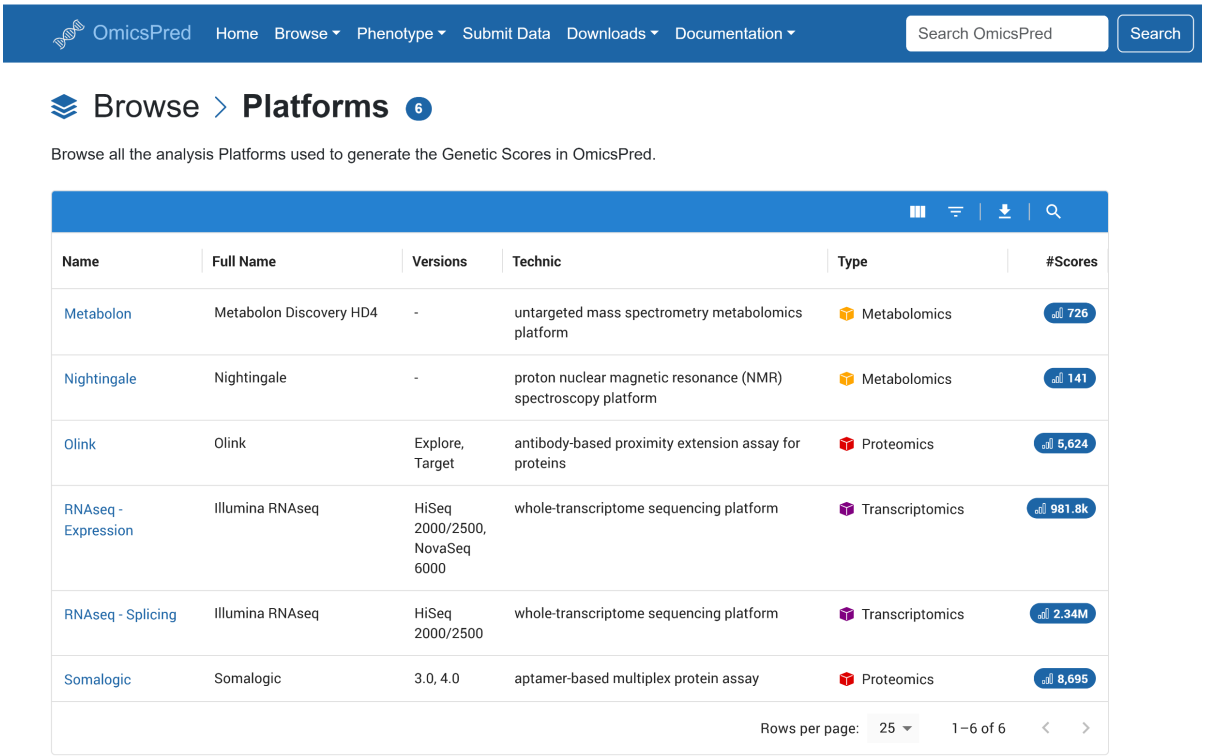

OmicsPred Home Browse Phenotype Submit Data Downloads Documentation Search OmicsPred Search

Browse > Platforms 6

Browse all the analysis Platforms used to generate the Genetic Scores in OmicsPred.

| Name | Full Name | Versions | Technic | Type | #Scores |
|---|---|---|---|---|---|
| Metabolon | Metabolon Discovery HD4 | - | untargeted mass spectrometry metabolomics platform | Metabolomics | 726 |
| Nightingale | Nightingale | - | proton nuclear magnetic resonance (NMR) spectroscopy platform | Metabolomics | 141 |
| Olink | Olink | Explore, Target | antibody-based proximity extension assay for proteins | Proteomics | 5,624 |
| RNAseq - Expression | Illumina RNAseq | HiSeq 2000/2500, NovaSeq 6000 | whole-transcriptome sequencing platform | Transcriptomics | 981.8k |
| RNAseq - Splicing | Illumina RNAseq | HiSeq 2000/2500 | whole-transcriptome sequencing platform | Transcriptomics | 2.34M |
| Somalogic | Somalogic | 3.0, 4.0 | aptamer-based multiplex protein assay | Proteomics | 8,695 |

Rows per page: 25 1–6 of 6

**Figure S3. Screenshot of the OmicsPred platforms page ([https://www.omicspred.org/platforms](https://www.omicspred.org/platforms)).** This page allows users to browse the list of omics platforms included in the catalog.

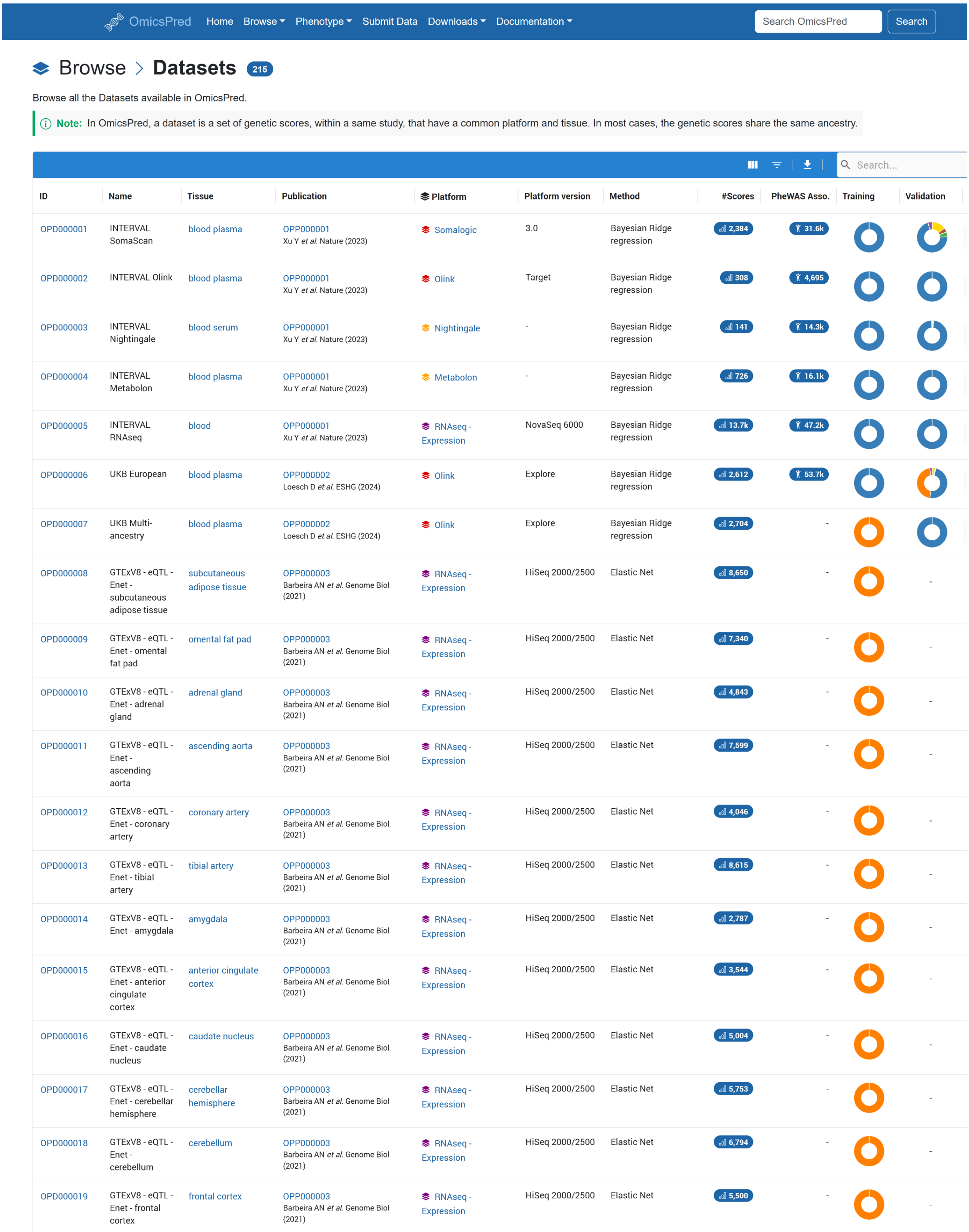

| ID | Name | Tissue | Publication | Platform | Platform version | Method | #Scores | PheWAS Asso. | Training | Validation |
|---|---|---|---|---|---|---|---|---|---|---|
| OPD000001 | INTERVAL SomaScan | blood plasma | OPP000001 Xu Y *et al.* Nature (2023) | Somalogic | 3.0 | Bayesian Ridge regression | 2,384 | 31.6k | | |
| OPD000002 | INTERVAL Olink | blood plasma | OPP000001 Xu Y *et al.* Nature (2023) | Olink | Target | Bayesian Ridge regression | 308 | 4,695 | | |
| OPD000003 | INTERVAL Nightingale | blood serum | OPP000001 Xu Y *et al.* Nature (2023) | Nightingale | - | Bayesian Ridge regression | 141 | 14.3k | | |
| OPD000004 | INTERVAL Metabolon | blood plasma | OPP000001 Xu Y *et al.* Nature (2023) | Metabolon | - | Bayesian Ridge regression | 726 | 16.1k | | |
| OPD000005 | INTERVAL RNAseq | blood | OPP000001 Xu Y *et al.* Nature (2023) | RNAseq - Expression | NovaSeq 6000 | Bayesian Ridge regression | 13.7k | 47.2k | | |
| OPD000006 | UKB European | blood plasma | OPP000002 Loesch D *et al.* ESHG (2024) | Olink | Explore | Bayesian Ridge regression | 2,612 | 53.7k | | |
| OPD000007 | UKB Multi-ancestry | blood plasma | OPP000002 Loesch D *et al.* ESHG (2024) | Olink | Explore | Bayesian Ridge regression | 2,704 | - | | |
| OPD000008 | GTExV8 - eQTL - Enet - subcutaneous adipose tissue | subcutaneous adipose tissue | OPP000003 Barbeira AN *et al.* Genome Biol (2021) | RNAseq - Expression | HiSeq 2000/2500 | Elastic Net | 8,650 | - | | - |
| OPD000009 | GTExV8 - eQTL - Enet - omental fat pad | omental fat pad | OPP000003 Barbeira AN *et al.* Genome Biol (2021) | RNAseq - Expression | HiSeq 2000/2500 | Elastic Net | 7,340 | - | | - |
| OPD000010 | GTExV8 - eQTL - Enet - adrenal gland | adrenal gland | OPP000003 Barbeira AN *et al.* Genome Biol (2021) | RNAseq - Expression | HiSeq 2000/2500 | Elastic Net | 4,843 | - | | - |
| OPD000011 | GTExV8 - eQTL - Enet - ascending aorta | ascending aorta | OPP000003 Barbeira AN *et al.* Genome Biol (2021) | RNAseq - Expression | HiSeq 2000/2500 | Elastic Net | 7,599 | - | | - |
| OPD000012 | GTExV8 - eQTL - Enet - coronary artery | coronary artery | OPP000003 Barbeira AN *et al.* Genome Biol (2021) | RNAseq - Expression | HiSeq 2000/2500 | Elastic Net | 4,046 | - | | - |
| OPD000013 | GTExV8 - eQTL - Enet - tibial artery | tibial artery | OPP000003 Barbeira AN *et al.* Genome Biol (2021) | RNAseq - Expression | HiSeq 2000/2500 | Elastic Net | 8,615 | - | | - |
| OPD000014 | GTExV8 - eQTL - Enet - amygdala | amygdala | OPP000003 Barbeira AN *et al.* Genome Biol (2021) | RNAseq - Expression | HiSeq 2000/2500 | Elastic Net | 2,787 | - | | - |
| OPD000015 | GTExV8 - eQTL - Enet - anterior cingulate cortex | anterior cingulate cortex | OPP000003 Barbeira AN *et al.* Genome Biol (2021) | RNAseq - Expression | HiSeq 2000/2500 | Elastic Net | 3,544 | - | | - |
| OPD000016 | GTExV8 - eQTL - Enet - caudate nucleus | caudate nucleus | OPP000003 Barbeira AN *et al.* Genome Biol (2021) | RNAseq - Expression | HiSeq 2000/2500 | Elastic Net | 5,004 | - | | - |
| OPD000017 | GTExV8 - eQTL - Enet - cerebellar hemisphere | cerebellar hemisphere | OPP000003 Barbeira AN *et al.* Genome Biol (2021) | RNAseq - Expression | HiSeq 2000/2500 | Elastic Net | 5,753 | - | | - |
| OPD000018 | GTExV8 - eQTL - Enet - cerebellum | cerebellum | OPP000003 Barbeira AN *et al.* Genome Biol (2021) | RNAseq - Expression | HiSeq 2000/2500 | Elastic Net | 6,794 | - | | - |
| OPD000019 | GTExV8 - eQTL - Enet - frontal cortex | frontal cortex | OPP000003 Barbeira AN *et al.* Genome Biol (2021) | RNAseq - Expression | HiSeq 2000/2500 | Elastic Net | 5,500 | - | | - |

**Figure S4. Screenshot of the OmicsPred datasets page (https://www.omicspred.org/datasets).** This page allows users to browse the list of datasets (groups of scores derived from a single publication that share a common platform, tissue, and training methodology) included in the catalog.

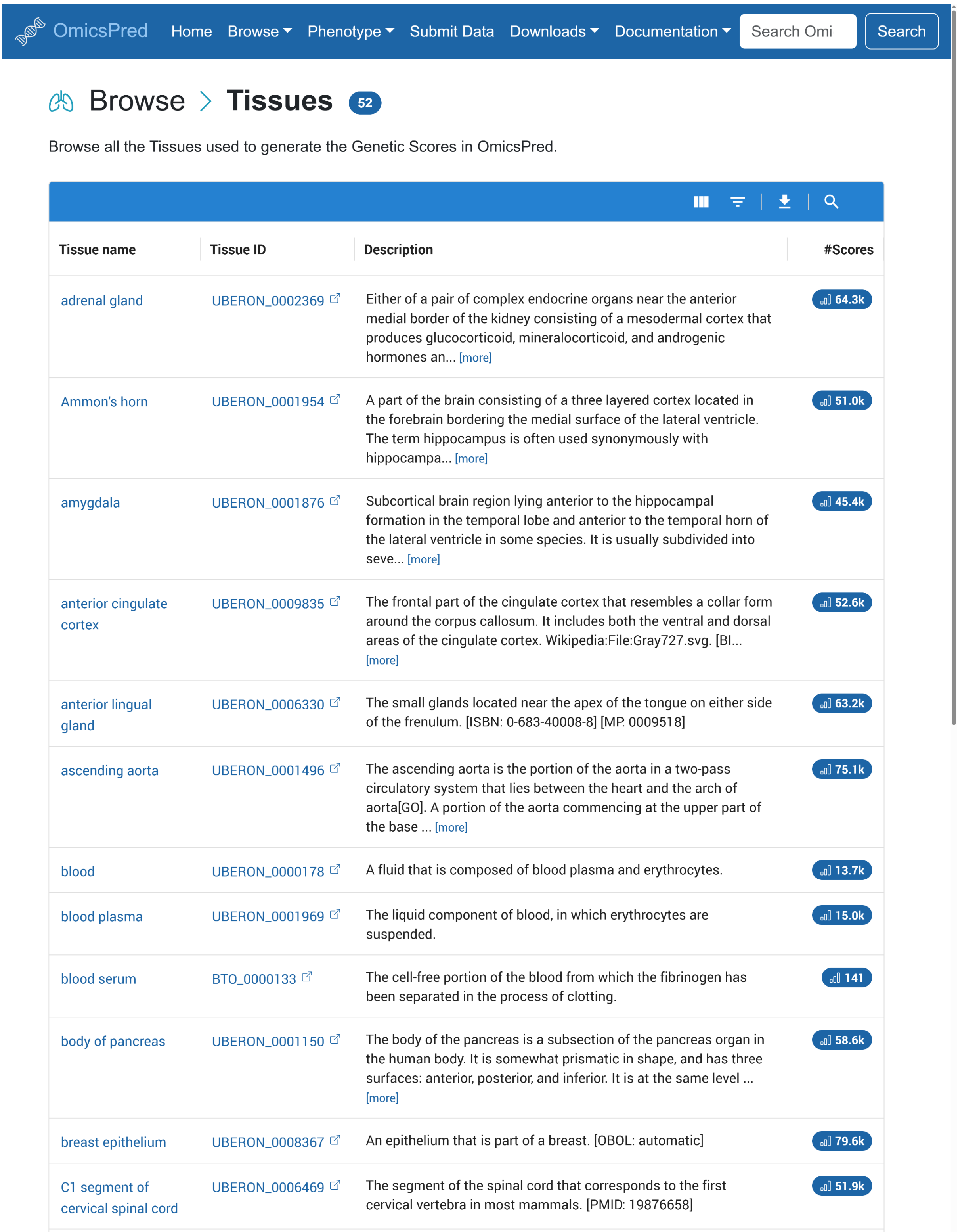

OmicsPred Home Browse Phenotype Submit Data Downloads Documentation Search Omi Search

Browse > Tissues 52

Browse all the Tissues used to generate the Genetic Scores in OmicsPred.

| Tissue name | Tissue ID | Description | #Scores |
|---|---|---|---|
| adrenal gland | UBERON_0002369 | Either of a pair of complex endocrine organs near the anterior medial border of the kidney consisting of a mesodermal cortex that produces glucocorticoid, mineralocorticoid, and androgenic hormones an... [more] | 64.3k |
| Ammon's horn | UBERON_0001954 | A part of the brain consisting of a three layered cortex located in the forebrain bordering the medial surface of the lateral ventricle. The term hippocampus is often used synonymously with hippocampa... [more] | 51.0k |
| amygdala | UBERON_0001876 | Subcortical brain region lying anterior to the hippocampal formation in the temporal lobe and anterior to the temporal horn of the lateral ventricle in some species. It is usually subdivided into seve... [more] | 45.4k |
| anterior cingulate cortex | UBERON_0009835 | The frontal part of the cingulate cortex that resembles a collar form around the corpus callosum. It includes both the ventral and dorsal areas of the cingulate cortex. Wikipedia:File:Gray727.svg. [Bl... [more] | 52.6k |
| anterior lingual gland | UBERON_0006330 | The small glands located near the apex of the tongue on either side of the frenulum. [ISBN: 0-683-40008-8] [MP. 0009518] | 63.2k |
| ascending aorta | UBERON_0001496 | The ascending aorta is the portion of the aorta in a two-pass circulatory system that lies between the heart and the arch of aorta[GO]. A portion of the aorta commencing at the upper part of the base ... [more] | 75.1k |
| blood | UBERON_0000178 | A fluid that is composed of blood plasma and erythrocytes. | 13.7k |
| blood plasma | UBERON_0001969 | The liquid component of blood, in which erythrocytes are suspended. | 15.0k |
| blood serum | BTO_0000133 | The cell-free portion of the blood from which the fibrinogen has been separated in the process of clotting. | 141 |
| body of pancreas | UBERON_0001150 | The body of the pancreas is a subsection of the pancreas organ in the human body. It is somewhat prismatic in shape, and has three surfaces: anterior, posterior, and inferior. It is at the same level ... [more] | 58.6k |
| breast epithelium | UBERON_0008367 | An epithelium that is part of a breast. [OBOL: automatic] | 79.6k |
| C1 segment of cervical spinal cord | UBERON_0006469 | The segment of the spinal cord that corresponds to the first cervical vertebra in most mammals. [PMID: 19876658] | 51.9k |

**Figure S5. Screenshot of the OmicsPred Tissues page (https://www.omicspred.org/tissues).** This page allows users to browse the list of tissues included in the catalog, important for navigating to individual cell/tissue-type pages necessary for identifying context-specific genetic predictors.

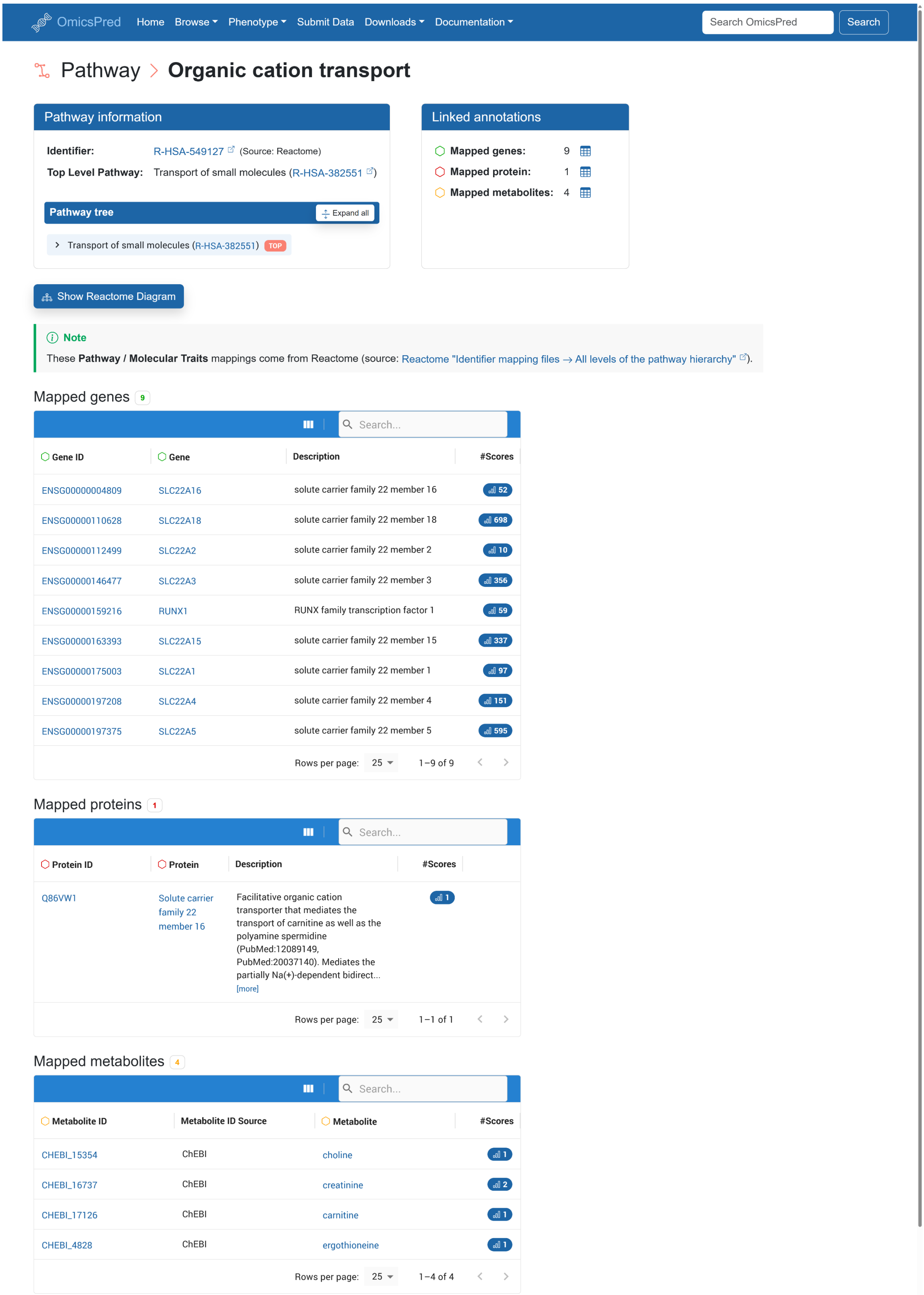

**Figure S5. Screenshot of an OmicsPred Pathway page (https://www.omicspred.org/pathway/R-HSA-549127).** Example of an OmicsPred pathway page displaying the scores that are mapped to a specific Reactome pathway (R-HSA-549127 in this example). Pathway-level mapping provides key biological context, facilitating functional interpretation and integration between different datasets.

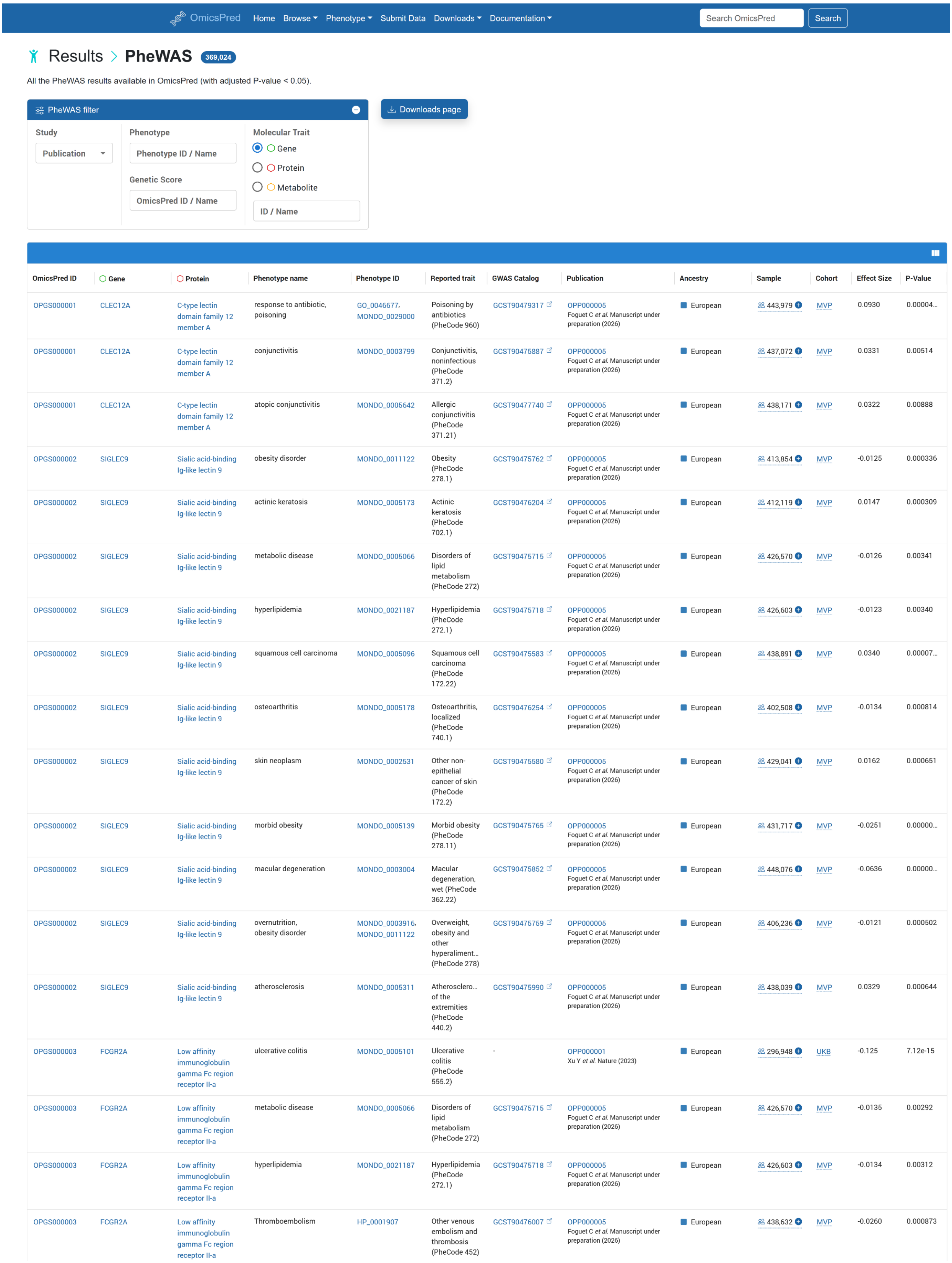

OmicsPred Home Browse Phenotype Submit Data Downloads Documentation Search OmicsPred Search

Results > PheWAS 369,024

All the PheWAS results available in OmicsPred (with adjusted P-value < 0.05).

PheWAS filter

Downloads page

Study: Publication

Phenotype: Phenotype ID / Name

Genetic Score: OmicsPred ID / Name

Molecular Trait: Gene / Protein / Metabolite

ID / Name

| OmicsPred ID | Gene | Protein | Phenotype name | Phenotype ID | Reported trait | GWAS Catalog | Publication | Ancestry | Sample | Cohort | Effect Size | P-Value |
|---|---|---|---|---|---|---|---|---|---|---|---|---|
| OPGS000001 | CLEC12A | C-type lectin domain family 12 member A | response to antibiotic, poisoning | GO_0046677, MONDO_0029000 | Poisoning by antibiotics (PheCode 960) | GCST90479317 | OPP000005 Foguet C *et al.* Manuscript under preparation (2026) | European | 443,979 | MVP | 0.0930 | 0.00004… |
| OPGS000001 | CLEC12A | C-type lectin domain family 12 member A | conjunctivitis | MONDO_0003799 | Conjunctivitis, noninfectious (PheCode 371.2) | GCST90475887 | OPP000005 Foguet C *et al.* Manuscript under preparation (2026) | European | 437,072 | MVP | 0.0331 | 0.00514 |
| OPGS000001 | CLEC12A | C-type lectin domain family 12 member A | atopic conjunctivitis | MONDO_0005642 | Allergic conjunctivitis (PheCode 371.21) | GCST90477740 | OPP000005 Foguet C *et al.* Manuscript under preparation (2026) | European | 438,171 | MVP | 0.0322 | 0.00888 |
| OPGS000002 | SIGLEC9 | Sialic acid-binding Ig-like lectin 9 | obesity disorder | MONDO_0011122 | Obesity (PheCode 278.1) | GCST90475762 | OPP000005 Foguet C *et al.* Manuscript under preparation (2026) | European | 413,854 | MVP | -0.0125 | 0.000336 |
| OPGS000002 | SIGLEC9 | Sialic acid-binding Ig-like lectin 9 | actinic keratosis | MONDO_0005173 | Actinic keratosis (PheCode 702.1) | GCST90476204 | OPP000005 Foguet C *et al.* Manuscript under preparation (2026) | European | 412,119 | MVP | 0.0147 | 0.000309 |
| OPGS000002 | SIGLEC9 | Sialic acid-binding Ig-like lectin 9 | metabolic disease | MONDO_0005066 | Disorders of lipid metabolism (PheCode 272) | GCST90475715 | OPP000005 Foguet C *et al.* Manuscript under preparation (2026) | European | 426,570 | MVP | -0.0126 | 0.00341 |
| OPGS000002 | SIGLEC9 | Sialic acid-binding Ig-like lectin 9 | hyperlipidemia | MONDO_0021187 | Hyperlipidemia (PheCode 272.1) | GCST90475718 | OPP000005 Foguet C *et al.* Manuscript under preparation (2026) | European | 426,603 | MVP | -0.0123 | 0.00340 |
| OPGS000002 | SIGLEC9 | Sialic acid-binding Ig-like lectin 9 | squamous cell carcinoma | MONDO_0005096 | Squamous cell carcinoma (PheCode 172.22) | GCST90475583 | OPP000005 Foguet C *et al.* Manuscript under preparation (2026) | European | 438,891 | MVP | 0.0340 | 0.00007… |
| OPGS000002 | SIGLEC9 | Sialic acid-binding Ig-like lectin 9 | osteoarthritis | MONDO_0005178 | Osteoarthritis, localized (PheCode 740.1) | GCST90476254 | OPP000005 Foguet C *et al.* Manuscript under preparation (2026) | European | 402,508 | MVP | -0.0134 | 0.000814 |
| OPGS000002 | SIGLEC9 | Sialic acid-binding Ig-like lectin 9 | skin neoplasm | MONDO_0002531 | Other non-epithelial cancer of skin (PheCode 172.2) | GCST90475580 | OPP000005 Foguet C *et al.* Manuscript under preparation (2026) | European | 429,041 | MVP | 0.0162 | 0.000651 |
| OPGS000002 | SIGLEC9 | Sialic acid-binding Ig-like lectin 9 | morbid obesity | MONDO_0005139 | Morbid obesity (PheCode 278.11) | GCST90475765 | OPP000005 Foguet C *et al.* Manuscript under preparation (2026) | European | 431,717 | MVP | -0.0251 | 0.00000… |
| OPGS000002 | SIGLEC9 | Sialic acid-binding Ig-like lectin 9 | macular degeneration | MONDO_0003004 | Macular degeneration, wet (PheCode 362.22) | GCST90475852 | OPP000005 Foguet C *et al.* Manuscript under preparation (2026) | European | 448,076 | MVP | -0.0636 | 0.00000… |
| OPGS000002 | SIGLEC9 | Sialic acid-binding Ig-like lectin 9 | overnutrition, obesity disorder | MONDO_0003916, MONDO_0011122 | Overweight, obesity and other hyperaliment… (PheCode 278) | GCST90475759 | OPP000005 Foguet C *et al.* Manuscript under preparation (2026) | European | 406,236 | MVP | -0.0121 | 0.000502 |
| OPGS000002 | SIGLEC9 | Sialic acid-binding Ig-like lectin 9 | atherosclerosis | MONDO_0005311 | Atherosclero… of the extremities (PheCode 440.2) | GCST90475990 | OPP000005 Foguet C *et al.* Manuscript under preparation (2026) | European | 438,039 | MVP | 0.0329 | 0.000644 |
| OPGS000003 | FCGR2A | Low affinity immunoglobulin gamma Fc region receptor II-a | ulcerative colitis | MONDO_0005101 | Ulcerative colitis (PheCode 555.2) | - | OPP000001 Xu Y *et al.* Nature (2023) | European | 296,948 | UKB | -0.125 | 7.12e-15 |
| OPGS000003 | FCGR2A | Low affinity immunoglobulin gamma Fc region receptor II-a | metabolic disease | MONDO_0005066 | Disorders of lipid metabolism (PheCode 272) | GCST90475715 | OPP000005 Foguet C *et al.* Manuscript under preparation (2026) | European | 426,570 | MVP | -0.0135 | 0.00292 |
| OPGS000003 | FCGR2A | Low affinity immunoglobulin gamma Fc region receptor II-a | hyperlipidemia | MONDO_0021187 | Hyperlipidemia (PheCode 272.1) | GCST90475718 | OPP000005 Foguet C *et al.* Manuscript under preparation (2026) | European | 426,603 | MVP | -0.0134 | 0.00312 |
| OPGS000003 | FCGR2A | Low affinity immunoglobulin gamma Fc region receptor II-a | Thromboembolism | HP_0001907 | Other venous embolism and thrombosis (PheCode 452) | GCST90476007 | OPP000005 Foguet C *et al.* Manuscript under preparation (2026) | European | 438,632 | MVP | -0.0260 | 0.000873 |

**Figure S6. Screenshot of the OmicsPred PheWAS page** (https://www.omicspred.org/phewas). This page allows users to browse the associations between genetically predicted molecular traits and disease outcomes, useful for hypothesis generation.
